# Prospective Bidirectional Relations Between Depression and Metabolic Health: 30 Year Follow-up from the NHLBI CARDIA Study

**DOI:** 10.1101/2023.03.08.23286983

**Authors:** Nicholas R. Moorehead, Jeffrey L. Goodie, David S. Krantz

**Author notes:** <u>Corresponding Author</u>: Nicholas R. Moorehead, 59^th^ Medical Operations Group, Wilford Hall Ambulatory Surgical Center, U.S. Air Force, Joint Base San Antonio – Lackland, TX 78236. <u>Note</u>: The opinions and assertions expressed herein are those of the authors and are not necessarily those of the Uniformed Services University or the U.S. Department of Defense. This article is based on a doctoral dissertation conducted by Nicholas R. Moorehead under the direction of David S. Krantz.

## Abstract

**Objective:** This study investigated prospective bidirectional relationships between depression and metabolic syndrome (MetS), and the moderating effects of race, sex, and health behaviors in a diverse cohort followed for 30 years.

**Methods:** Data were analyzed from the NHLBI CARDIA study, a 30 year-prospective study of young adults (*N* = 5113; *M* age = 24.76 (*SD* = 3.63) at baseline; 45% male) who were tested every 5 years between 1985-2015. Measures included biological assessments of MetS components, and self-reported depressive symptoms based on the Center for Epidemiologic Studies Depression (CESD) scale. Data analyses included bi-directional general estimating equations analyses of time-lagged associations between depressive symptoms and MetS.

**Results:** There was a consistent, bi-directional relationship between depressive symptoms and MetS over time. Individuals with more CESD depressive symptoms were more likely to develop MetS over time compared to those reporting fewer symptoms (Wald Chi-Square = 7.09 (1), *p <* 0.008), and MetS was similarly predictive of CESD. MetS more consistently predicted depressive symptoms at each 5-year exam than depressive symptoms predicted MetS. Race and sex moderated relationships between depression and MetS, with White females, White individuals overall, and females overall demonstrating significant relationships. Health behaviors were not related to depression-MetS associations.

**Conclusion:** In a diverse young adult population prospectively followed into late middle age, MetS more consistently predicted depression over time than depression predicted MetS. The relation between MetS and depressive symptoms was moderated by race and sex, but not health behaviors.

---

Metabolic syndrome (MetS), defined by the presence of obesity, hyperglycemia, dyslipidemia, and hypertension, is an important risk factor for the development of cardiovascular disease (CVD; Huang, 2009). Research has also shown that MetS is associated with depression (Akbaraly et al., 2009; Goldbacher et al., 2009; Pan et al., 2012), and depression is also a risk factor for the development of CVD (Ogunmoroti et al., 2022). Further, it has been suggested that the relationship between depression and MetS is bidirectional, with both depression preceding the onset of MetS and MetS predicting the onset of depression (Pan et al., 2012). Research involving multiple longitudinal assessments of depression and MetS in the same sample is needed to test directional associations between depression and MetS.

## Relationships between Depressive Symptoms and MetS

The predictive relationship of depression for metabolic syndrome is well-established. For example, a recent meta-analysis of 49 studies (Moradi et al., 2021) with a total sample size of nearly 400,000, observed a greater likelihood of developing MetS among depressed individuals compared to those who were non-depressed. These relationships held for both cross-sectional and cohort studies, and there was evidence of a stronger relationship between depression and MetS in women, compared to men. Evidence further indicates that associations between depression and MetS may differ by race as well as gender (Womack et al., 2016). However, because the incidence of MetS increases with age, research is needed to determine whether there are gender and race differences in the relationships between depression and MetS over the lifespan (Moradi et al., 2021; Pan et al., 2012; Womack et al., 2016).

Mechanisms suggested for a causal relationship of depressive symptoms to MetS include associations between depression and biological and behavioral changes that promote the development of MetS. Neuroendocrine, autonomic, and inflammatory effects of depression can have effects on factors contributing to MetS such as abdominal fat, metabolism, and blood pressure (Akbaraly et al., 2009; Hryhorczuk et al., 2013; Liaw et al., 2015; Pan et al., 2012).

Depression is also associated with behavioral risk factors that increase the development of MetS. These include reduced exercise (Belvederi Murri et al., 2019), obesity (Hryhorczuk et al., 2013; Luppino et al., 2010; Needham et al., 2010), poor diet (Ljungberg et al., 2020), smoking (Bakhshaie et al., 2015; Fluharty et al., 2017; Mathew et al., 2017), sleep (Murphy & Peterson, 2015), and reduced medication compliance and health behaviors (Akbaraly et al., 2009; Grenard et al., 2011; Liaw et al., 2015; Womack et al., 2016).

## Bi-directional Relationships Between Depression and MetS

The overlap between physiological, social, behavioral, and emotional sequalae of both MetS and depression has led researchers to investigate the extent to which this relationship may be bi-directional (i.e., the presence of metabolic syndrome may be predictive of depression as well). However, evidence for metabolic syndrome as a predictor of depression is equivocal, with both positive (Akbaraly et al., 2009), and negative findings (Foley et al., 2010). Summarizing this literature, a meta-analysis of 29 cross-sectional and 11 cohort studies of the relation of depression and MetS included 11 studies that used MetS as a predictor of depression and 12 studies examining depression as a predictor of MetS (Pan et al, 2012). These authors concluded that the aggregate data supported a bidirectional relationship between depression and MetS. The relation between depression and MetS appeared to be stronger in cross-sectional studies that measured depression using self-reported symptom scales compared to clinical interviews and formal clinical diagnosis (Pan et al., 2012).

There are plausible mechanisms that may account for a possible predictive relationship of MetS for depression. MetS is associated with increased levels of inflammatory cytokines such as C-reaction protein, interleukin, as well as leptin resistance which are also associated with depressed mood (Akbaraly et al., 2009). Behavioral factors that are associated with MetS (e.g., sedentary behavior, poor diet, obesity, and sleep disturbance) might increase an individual’s likelihood of developing depression as a reduction in behavioral activation can lead to increased negative thoughts and emotions (Akbaraly et al., 2009; Gozal et al., 2016; Hryhorczuk et al., 2013; Liaw et al., 2015; Pan et al., 2012). In addition, many of the treatments recommended for MetS (e.g., physical activity, eat a healthy diet, improve sleep) are similar to behavioral treatments effective in reducing depression (Hiles et al., 2016; Pan et al., 2012).

## Relations of Sex, Age, and Race with Depression and Metabolic Syndrome

Demographic factors such as sex, age, and race are associated with the incidence and prevalence of both depression and metabolic syndrome (Bailey et al., 2019; Gurka et al., 2014; Hargrove et al., 2020; Womack et al., 2016). Women experience twice the risk of depression when compared to men (Kessler & Bromet, 2013; Sutin et al., 2013). With regard to age, longitudinal research indicates that depressive symptoms are highest in young adulthood, decrease in middle age, and subsequently increase again in older adults (Sutin et al, 2013). Metabolic syndrome increases in prevalence with age, with the age-related increase being greater in women across the lifespan (Vishram et al., 2014). However, it is not known whether there are bi-directional relationships between depression and metabolic syndrome across the lifespan for extended periods from young adulthood to older ages (Pan et al., 2012). Reviews and meta-analyses of multiple studies have been able to indirectly assess the presence of a bidirectional relation between depression and MetS, but little is known about a possible bidirectional relation in the same individuals (Moradi et al., 2021; Pan et al., 2012; Womack et al., 2016).

A recent report from the NHLBI CARDIA study examined relationships between MetS and depression utilizing 15-year follow-up (Womack et al., 2016). The ages at follow-up in that study were between 33 and 45 years. The prevalence of MetS was highest among Black women, followed by White men, Black men, and White women. As in prior research (Womack et al., 2016), incidence of MetS was higher for those who reported more depressive symptoms. In addition, depressive symptoms were more strongly associated with the development of MetS in White men and White women, with weak or no associations among Black men and women (Womack et al., 2016).

## Current Study

The presence, strength, and consistency of a possible bi-directional relationship over the lifespan is not well understood. Therefore, the primary goal of the present study is to examine possible bi-directional relationships in a 30-year longitudinal follow-up with assessments made every 5 years in the same sample of participants. In addition, this study also extended the findings of Womack et al. (2016) in the CARDIA study, using a longer follow-up (30 vs. 15 years) from young adulthood (ages 18-30) to late middle age (ages 48 - 60). Another study goal was to determine whether health behaviors and demographic factors influenced these relationships. Finally, we further extended CARDIA findings by examining these relationships among Black and White men and women using this longer follow-up. We hypothesized that there would be a bidirectional association between metabolic health and depression, and that race, sex, and health behaviors would moderate the relation between depression and metabolic health.

## METHODS

This study utilized data from the limited access dataset from National Heart, Lung, and Blood Institute (NHLBI) Coronary Artery Disease in Young Adults (CARDIA) study (NHLBI, 2021) of a diverse population of young adults followed up for 30 years between 1985 and 2015. Participants in were 5115 Black (*N* = 2637) and White (*N* = 2462) men (*N* = 2328) and women (*N* = 2785) age 18-30 at intake who were re-tested at follow-ups during 1987 (Year 2), 1990 (Year 5), 1992 (Year 7), 1995 (Year 10), 2000 (Year 15), 2005 (Year 20), 2010 (Year 25), and most recently 2015 (Year 30). Detailed methodology for CARDIA is presented elsewhere (Friedman et al., 1988). Due to the low number of individuals identifying as Hispanic (*N* = 14), these individuals were excluded from analyses. The present analyses use assessments from 1990 (Year 5), 1995 (Year 10), 2000 (Year 15), 2005 (Year 20), 2010 (Year 25), and 2015 (Year 30).

### Measures

#### Demographic and Clinical Data

At baseline and every follow-up, medical and demographic measures were collected. Variables of interest for the present analyses included: anthropomorphic and sociodemographic variables such as body mass index, age, race, and sex; smoking status; alcohol consumption; depression/depressive symptoms; and biological variables including MetS and its components.

#### Physical Activity

To measure physical activity, we utilized the CARDIA self-report physical activity history measured at each examination (Camhi et al., 2013; Jacobs et al., 1989). This questionnaire measure was developed specifically for use in the CARDIA study to provide a suitable reliable and valid brief, yet comprehensive, questionnaire to measure different types of physical activity (Camhi et al., 2013; Jacobs et al., 1989). Questions pertain to the types of physical activity, perceived level of difficulty/rigor, and the total amount in hours/minutes at either daily or weekly intervals. A physical activity score based on the time, intensity, and frequency of each documented activity was calculated. Total hours per week of moderate, hard, very hard activity are also derived from the physical activity recall.

#### Center for Epidemiological Studies Depression Scale (CESD)

This 20-item questionnaire is widely used to assess depression and depressive disorder in population studies. CESD items consist of Likert scales ranging from “rarely or none of the time” to “most or all of the time,” with scale scores ranging from 0 to 60. Although not intended for clinical diagnosis, a score ≥16 has been associated with suspected depression diagnosis (Radloff, 1977). The CESD has good sensitivity and specificity for detecting depression, is useful across the lifespan, and is reliable when compared to other depression scales (Cosco et al., 2017; Radloff, 1977; Shafer, 2006; Vilagut et al., 2016). In CARDIA, The CESD was administered at baseline and at follow-up years 5, 10, 15, 20, 25, and 30. Therefore, the final depression measure was 30 years following intake, and the first measure of depression used was at year 5.

#### Bloodwork and Identification of MetS

Before each collection of blood, participants fasted for at least 12 hours in accordance with standardized protocols (Friedman et al., 1988). At each exam, participants had their blood pressure taken three times with the average of the final two being recorded. Waist circumference was determined by the average of two waist circumference measurements (Friedman et al., 1988; Womack et al., 2016).

For this study, we used the NCEP-ATP III criteria to define MetS (Grundy, Brewer, et al., 2004; Grundy, Hansen, et al., 2004). This criterion indicates that no single variable threshold is required for MetS, but subjects must meet 3 of the 5 following criteria: (1) Obesity defined by a waist circumference greater than 40 in. (101.6 cm) for men or greater than 35 in. (88.9 cm) for women; (2) Hyperglycemia indicated by fasting glucose greater than or equal to 100 mg/dl or by a prescription; (3) Dyslipidemia indicated by any of the following: triglyceride levels ≥ 150 mg/dl; medication treatment for dyslipidemia; HDL-C < 40 mg/dl for men and < 50 mg/dl for women (4) Hypertension indicated by systolic blood pressure >130 mmHg, diastolic blood pressure >85 mmHg, or use of prescribed antihypertensive medications. MetS was coded as a binary variable because the presence of MetS is considered to be a diagnostic category without an established criteria for severity of the syndrome (Grundy, Brewer, et al., 2004; Grundy, Hansen, et al., 2004; Huang, 2009).

### Data analysis

#### Relations Between Depression and MetS

To determine longitudinal relationships between the predictor variable depression to the MetS outcome over time, and the moderating effects of health behaviors on that relationship, data were analyzed using general estimating equations (GEE) in SPSS. This analysis allowed for both a cross-sectional and longitudinal output in a single GEE model accounting for fixed effects. This means a single analysis provided between-groups (between-subjects) relationships at time points baseline, Year 5, Year 10, Year 15, Year 20, Year 25, and Year 30, longitudinal (within-subjects) results over the entire study period, and allowed for pairwise comparisons to be included to compare groups at various time points. The GEE approach also has the benefit of being robust enough to include participants even if data are missing at one or several timepoints, and significantly reduce likelihood of Type 1 error by reducing the total numbers of analyses/models conducted.

Depression was coded as binary (CESD < 16 = “not depressed”; CESD ≥ 16 = “depressed”). MetS was coded as binary (MetS = ≥3 criterion variables [i.e., hypertension, obesity, dyslipidemia by triglycerides, dyslipidemia by HDL-C, or hyperglycemia]; vs No MetS (<3 or more criterion variables) at each time point. This allowed determination of an odds ratio (OR) for developing MetS relative to the reference (not depressed) group. Covariates (age, sex, race, physical activity, smoking, alcohol consumption, and time) were included in the analyses.

#### Bi-directional Relations Between Depression and MetS

To examine bi-directional associations between depression and MetS, two sets of lagged time-series logistic regressions were utilized. First, to determine if depression predicts MetS, binary coding of depression at each exam year served as the predictor variable and MetS at the subsequent exam year was the outcome variable. Five logistic regressions were conducted: depression at Year 5, 10, 15, 20, and 25 predicting MetS status at Year 10, 15, 20, 25, and 30, respectively. To determine whether depression predicted future MetS, 5 logistic regressions were also conducted with depression at each exam year used to predict MetS status at the next exam year. Covariates (age, sex, race, physical activity, smoking, alcohol consumption, and time) were included in the analyses.

#### Depression and MetS Relations with Sex and Race

To examine whether relationships between depression and MetS outcomes differ by sex (male vs. female) and race (Black vs. White), Depression and MetS were again coded as binary variables, and sex or race were included as possible moderators of relationships between depression and MetS. Three GEE models were conducted, one for testing the possible moderating effects of sex, one for testing the moderating effects of race, and on for testing the moderating effects of race and sex together.

## RESULTS

### Variables

#### Demographics

As shown in Table 1, the baseline sample had an approximately even racial Black/White split (51% identifying as Black), and an even male/female split (55% females). Over the course of the study the total number of participants decreased from 5113 at baseline to 3357 at Year 30 demonstrating a 66% retention rate. At Year 30, the split across racial and sex lines differed (57% females), and 48% Black.

**Table 1.** Demographics

| Study Exam Year | Sex |  | Race |  |  | Mean Age (Std Dev) | N | % Retention |
| --- | --- | --- | --- | --- | --- | --- | --- | --- |
|  | Male | Female | Hispanic | Black | White |  |  |  |
| Baseline | 2328<br>(45.5%) | 2785<br>(54.5%) | 14<br>(0.3%) | 2637<br>(51.6%) | 2462<br>(48.2%) | 24.76<br>(3.63) | 5113 | 100% |
| Yr5 | 1959<br>(45.0%) | 2392<br>(55.0%) | 13<br>(0.3%) | 2119<br>(48.7%) | 2219<br>(51.0%) | 29.90<br>(3.60) | 4351 | 85% |
| Yr10 | 1755<br>(44.5%) | 2188<br>(55.5%) | 13<br>(0.3%) | 1922<br>(48.7%) | 2008<br>(50.9%) | 34.92<br>(3.61) | 3943 | 77% |
| Yr15 | 1619<br>(44.1%) | 2049<br>(55.9%) | 11<br>(0.3%) | 1729<br>(47.1%) | 1928<br>(52.6%) | 40.01<br>(3.60) | 3668 | 72% |
| Yr20 | 1535<br>(43.3%) | 2012<br>(56.7%) | 11<br>(0.3%) | 1651<br>(46.5%) | 1885<br>(53.1%) | 45.01<br>(3.59) | 3547 | 69% |
| Yr25 | 1517<br>(43.4%) | 1979<br>(56.6%) | 9<br>(0.3%) | 1639<br>(46.9%) | 1848<br>(52.9%) | 50.00<br>(3.59) | 3496 | 68% |
| Yr30 | 1444<br>(43.0%) | 1913<br>(57.0%) | 9<br>(0.3%) | 1605<br>(47.8%) | 1743<br>(51.9%) | 54.94<br>(3.57) | 3357 | 66% |

*Note.* Percentages are relative to the whole population (e.g., 2328 males at baseline represent 45.5% of the initial sample of 5113).

#### Health Behaviors

Those who reporting “never smoked” stayed relatively consistent, (56% at baseline vs. 63% of the retained sample at Year 30; see Table 2). Comparing baseline to Year 30: current smokers decreased (approximately 31% at baseline vs. 14%); reported alcohol consumed stayed relatively constant; and physical activity slightly declined.

**Table 2.** Health Behaviors

| Exam<br>Year | Alcohol Consumed<br>(ml/alcohol/ day) | Physical<br>Activity Score | BMI<br>($kg/m^2$) | Smoking | | |
| --- | --- | --- | --- | --- | --- | --- |
|  |  |  |  | Never | Not Currently | Yes |
| baseline | 12.07 (21.94) | 420.14 (300.77) | 24.44 (4.81) | 2893 (56.6%) | 653 (12.8%) | 1566 (30.6%) |
| Yr5 | 11.21 (25.61) | 379.42 (292.55) | 26.09 (5.62) | 2486 (57.3%) | 613 (14.1%) | 1242 (28.6%) |
| Yr10 | 10.94 (22.05) | 330.90 (274.93) | 27.46 (6.25) | 2275 (57.9%) | 648 (16.5%) | 1004 (25.6%) |
| Yr15 | 10.97 (24.91) | 347.28 (283.58) | 28.68 (6.49) | 2193 (59.9%) | 664 (18.1%) | 807 (22.0%) |
| Yr20 | 10.84 (22.24) | 335.87 (274.13) | 29.32 (6.53) | 2150 (61.2%) | 681 (19.4%) | 683 (19.4%) |
| Yr25 | 11.71 (23.44) | 337.79 (275.64) | 30.06 (6.76) | 2106 (61.1%) | 749 (21.7%) | 589 (17.1%) |
| Yr30 | 11.23 (19.82) | 321.34 (271.50) | 30.43 (6.78) | 2079 (62.8%) | 768 (23.2%) | 463 (14.0%) |

#### MetS

Table 3 presents rates of those meeting overall MetS diagnostic criteria, and rates with MetS components of hypertension, obesity, dyslipidemia, hyperglycemia; fasting glucose was not measured at Year 5. The number of positive MetS components, and number of individuals meeting criteria for MetS increases over the course of the study.

**Table 3.** CESD Continuous and Binary Depression Scores Based on CESD >16 and MetS Diagnoses

| Year | CESD (SD) | Depression<br>(no) | Depression<br>(yes) | MetS Dx |
| --- | --- | --- | --- | --- |
| Baseline | N/A | N/A | N/A | 106 (2.1%) |
| Yr5 | 11.24 (8.12) | 3258 (75.7%) | 1042 (24.3%) | 130 (0.3%) |
| Yr10 | 10.70 (8.22) | 3032 (78.2%) | 843 (21.8%) | 361 (9.2%) |
| Yr15 | 9.16 (7.85) | 3005 (83%) | 617 (17%) | 493 (13.4%) |
| Yr20 | 9.33 (7.88) | 2835 (82.2%) | 612 (17.8%) | 679 (19.1%) |
| Yr25 | 9.47 (7.74) | 2855 (82.7%) | 598 (17.3%) | 712 (20.4%) |
| Yr30 | 8.95 (7.96) | 2707 (82.9%) | 560 (17.1%) | 737 (22.0%) |

#### Depression

Table 3 summarizes depression data. Participants scoring in the depressed range was highest at Year 5 (i.e., 24% of the sample), dropped to approximately 17% of the sample at Year 15, and remained stable until Year 30. The CESD was not given used at baseline.

#### Relations Between Depression, MetS, and Health Behaviors

As summarized in Table 4, depression status was significantly related to MetS status (Wald Chi-Square = 7.09 (1), *p <* 0.008) over the course of the study. The effect of time by itself was significant (*p <* 0.0001). The time by depression interaction was not significant (*p =* 0.45), suggesting that the relation between depression and MetS diagnosis does not vary over time.

**Table 4.** Model Effects for relation between predictor variables and MetS (as a dependent variable)

| Variable | Wald |  |  |
| --- | --- | --- | --- |
|  | Chi-Square | df | *p-level* |
| (Intercept) | 119.703 | 1 | 0.000 |
| Time | 515.801 | 5 | 0.000 |
| Age | 18.277 | 1 | 0.000 |
| Sex | 38.622 | 1 | 0.000 |
| Race | 6.716 | 1 | 0.010 |
| Physical Activity | 105.615 | 1 | 0.000 |
| Smoking | 4.965 | 2 | 0.084 |
| Alcohol Consumption | 1.179 | 1 | 0.278 |
| Depression | 7.090 | 1 | 0.008 |
| Time by Depression<br>Interaction | 4.727 | 5 | 0.450 |

Physical activity, smoking, and alcohol consumption were each examined as possible moderators of the predictive relationships between depression and MetS. For CESD depression as the independent variable and MetS diagnosis as the dependent variable, in cross-sectional, longitudinal, and/or lagged-times series analyses, no individual health behavior had a significant moderating effect on the relationship between depression and MetS diagnosis. Although there were significant effects for depression and for physical activity on MetS in the overall model (*p* < 0.05), physical activity did not moderate these relationships. Therefore, the relationship to MetS to depression did not vary by physical activity or any other health behavior.

#### Bi-directional Relations Between Depression and MetS

Results for the first series of lagged logistic regressions examining whether depression predicts MetS status are presented in Figure 1A. Three of the 5 time-lagged results for depression as a predictor of MetS (years 10-15, 15-20, and 25-30) were significant (*p* < 0.05). The OR’s stay consistent across time, ranging diagnosis of depression at Year 10 preceding development of MetS at Year 15 (OR = 1.38, 95% CI [1.08, 1.75], *p* < 0.009), to (OR = 1.32, 95% CI [1.05, 1.66], *p* < 0.016) for depression at year 25 predicting MetS at Year 30.

**Figure 1.**
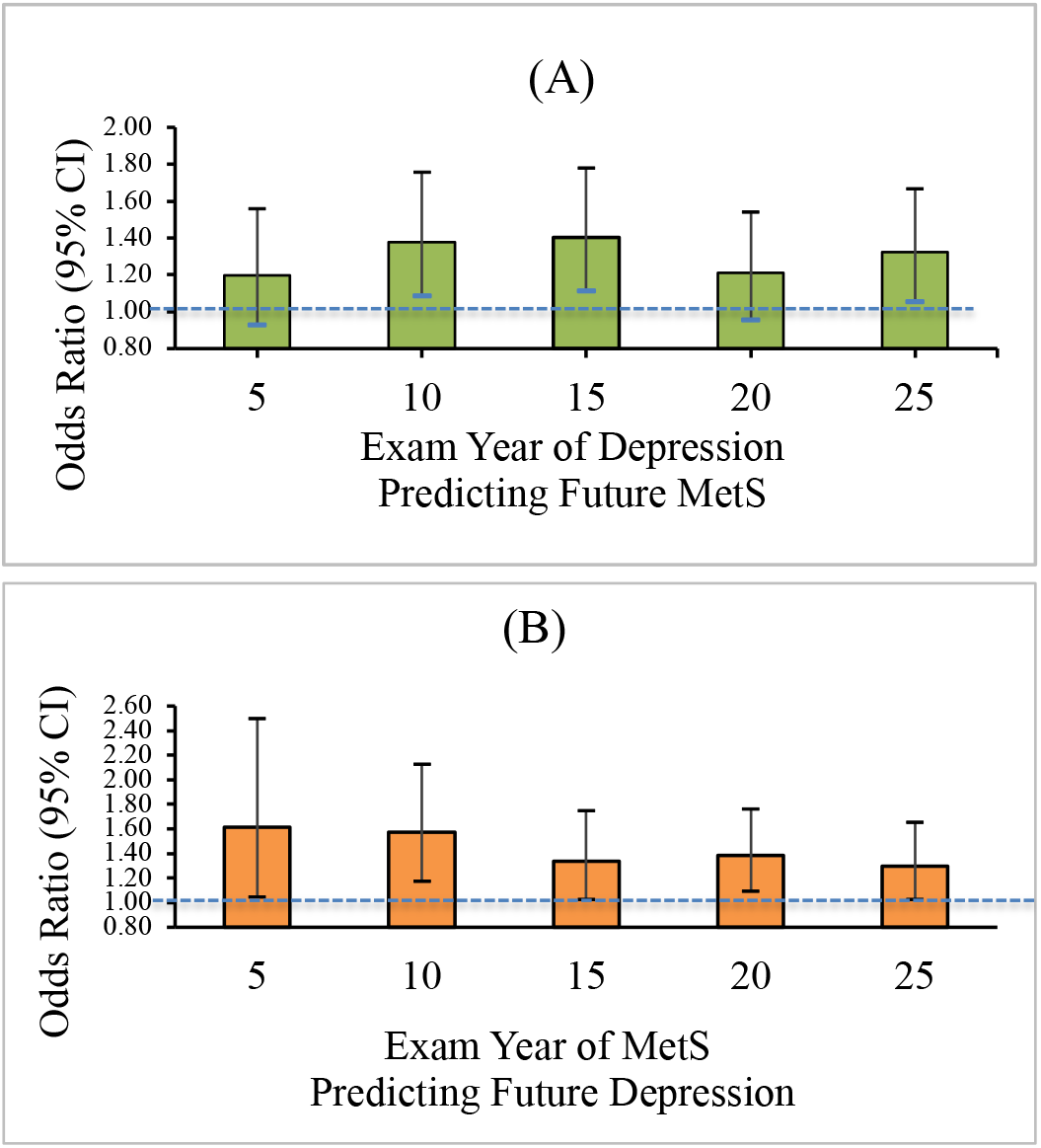
(A) Odds (95% CI) of Developing Metabolic Syndrome Predicted by Depression at Previous Exam 5-Years Earlier based on Lagged Time Series Analyses; (B) Odds (95% CI) of Developing Depression Predicted by Metabolic Syndrome at Pevious Exam 5-Years Earlier based on Results of Lagged Time Series Analyses *Note*. MetS=Metabolic Syndrome. In each analysis, the independent variable (presence/absence of either depression or MetS) was used to predict presence/absence of either MetS or depression, respectively, at the next 5-year exam. (A) Odds (95% CI) of depression (yes/no) prospectively predicting future presence of MetS (yes/no) at the subsequent exam year. *Years 10, 15, and 25 are significant at the *p< 0*.*05* level; (B) Odds (95% CI) of presence of MetS (yes/no) at each exam year prospectively predicting depression (yes/no) at the subsequent exam year. *All exam years are significant at the *p<0*.*05* level.

Figure 1B presents results for the series of lagged logistic regressions examining MetS as a predictor of depression. Although the relationships were significant at all time points, the OR’s were somewhat more variable in their magnitude than in analyses of depression predicting MetS. At Year 5, if a person was diagnosed with MetS vs. no MetS, they were 1.61 times more likely to have depression at Year 10 (OR = 1.61, 95% CI [1.05, 2.50], *p* < 0.030). For Year 25 MetS predicting Year 30 depression, the OR was 1.30, 95% CI [1.02. 1.65], *p* < 0.032.

#### Sex, Race, and Relationships between Depression and MetS

Table 5 presents results of three separate GEE models examining whether relationships between depression and MetS outcomes differ by race (Black vs. White), by sex (male vs. female), and their combination. In the first model, race did not have significant relation with MetS diagnosis; nor did it moderate the relation between depression and MetS status (*p* = 0.076). In the second model, Sex had a significant relation with MetS diagnosis (*p <* 0.001), and sex moderated the relation between depression and MetS diagnosis (*p* < 0.004), with effects stronger in women. In the third model, Sex and Race significantly interact (*p* < 0.001), and together moderate the relation between depression and MetS (*p* < .002). Figure 2 displays the ORs and 95% confidence intervals for comparative effects of the Sex and Race groups as possible moderators of depression as a predictor of MetS. As indicated in the figure, the only significant sex/race groups are White females, females, and White individuals.

**Table 5.** Results from three separate GEE models testing the moderating effects of Race, Sex, and the interaction of Race x Sex on the relationship between Depression (as the predictor) and Metabolic Syndrome (the outcome).

| Variable | Wald<br>Chi-Square | df | *p*-level |
| --- | --- | --- | --- |
| Race | 2.627 | 1 | 0.105 |
| Depression | 8.017 | 1 | 0.005 |
| Race x Depression<br>Interaction | 3.147 | 1 | 0.076 |
| Sex | 18.515 | 1 | 0.000 |
| Depression | 3.649 | 1 | 0.056 |
| Sex x Depression<br>Interaction | 8.383 | 1 | 0.004 |
| Sex x Race | 42.874 | 3 | 0.000 |
| Depression | 5.135 | 1 | 0.023 |
| Sex x Race x<br>Depression Interaction | 14.698 | 3 | 0.002 |

*Note.* The table presents the results of 3 separate GEE models of the interactions between race and sex on the relationship between Depression and Metabolic Syndrome (the outcome). From top to bottom, these models examine the following interactions: Race x Depression, Sex x Depression, and Sex x Race x Depression.

**Figure 2.**
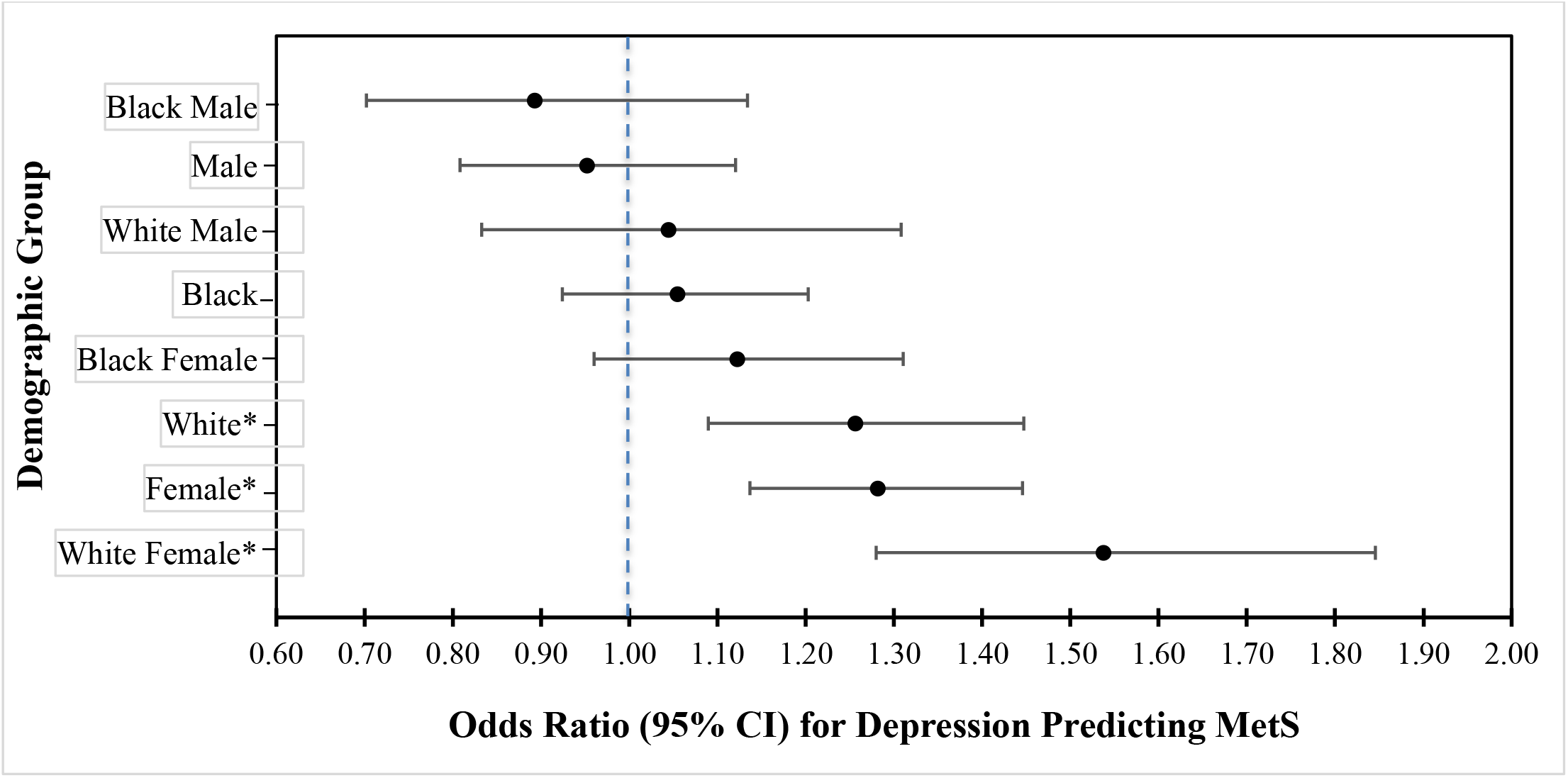
Summary Forest Plot of Odds Ratios (95% CI) for Moderating Effects of Sex and Race on Depression as a Predictor of MetS (see footnote) *Note*. Odds of having metabolic syndrome *if that demographic has depression relative to that demographic not having depression* (e.g., White Females with depression are 1.53 times more likely to have metabolic syndrome compared to White females without depression). Error bars represent the 95% CI for each odds ratio. *Indicates a significant relationship between Depression and MetS for that group.

## DISCUSSION

This study is unique in examining bi-directional, longitudinal relations between depression and MetS in a young adult cohort as they progressed to middle-age over a 30-year period. The present findings of significant relationships between depression and MetS over time are consistent with prior findings from several studies and meta-analyses (Akbaraly et al., 2009; Foley et al., 2010; Goldbacher et al., 2009; Pan et al., 2012). In addition, this study found bi-directional relationships between depression and MetS over the 30-year time period.

The present results suggest that weaker findings found in cross-sectional comparisons in prior studies may reflect stronger relationships between depression and MetS at certain points in the lifespan, and/or that cross-sectional assessments at only a single time point may contribute to weaker associations than longitudinal measurements at multiple time points. Interestingly, the present findings were obtained despite the fact that the NCEP ATP III criteria were used to define MetS in the present study. Pan et al. (2012) noted that in other studies using NCEP ATP III criteria to define MetS, the relation was found to be weaker, likely due to the NCEP ATP III requiring a lower threshold to meet diagnostic criteria compared to other diagnostic criteria for MetS (Pan et al., 2012). Also consistent with prior research are our findings over 30-years that the relation between MetS and depression is weak or not significant in males, and more consistently found in females (Pan et al., 2012). These findings are addressed later in the discussion section.

Prior studies did not assess bidirectional relationships between depression and MetS in the same study cohort. In this study, unique analyses using lagged time series models were made possible by the multiple assessments over time in CARDIA, and the predictive relation between depression and MetS was slightly stronger and more consistent for MetS prospectively predicting depression than the opposite (Akbaraly et al., 2009; Foley et al., 2010; Goldbacher et al., 2009; Hiles et al., 2016; Moradi et al., 2021; Pan et al., 2012; Womack et al., 2016). This finding of a more consistent effect for MetS predicting depression is supported by some studies (Akbaraly et al., 2009; Pan et al., 2012) but not others (Hiles et al., 2016). However, compared to prior studies not looking at both relationships in the same cohort, the present findings based on multiple longitudinal datapoints in the same cohort over 30 years are convincing. However, further study is needed to assess the consistency of these bidirectional relationships in other cohorts.

### Possible Mechanisms for Links Between Depression and MetS

Several causal mechanisms have been suggested to explain the predictive relation of depression for MetS; however, the predictive relationship of MetS for depression might seem less intuitive. Factors contributing to both directional relationships may include biological factors (e.g., elevated stress levels causing HPA axis dysregulation, changes in levels of cortisol and glucocorticoids, and increased pro-inflammatory responses), psychological factors (e.g., increased social isolation associated with depression), and behavioral changes (e.g., health-impairing behaviors such as smoking, alcoholism, decreased exercise levels, poor diet).

In the present study, moderating factors examined were physical activity, smoking, and alcohol consumption. Of these, only physical activity demonstrated a moderating effect on MetS. The reasons for null findings for smoking and alcohol consumption, which are known to increase risk of MetS (Boyle et al., 2018; Carroll et al., 2019; Sun et al., 2012), are not clear. Although previous reports indicate that neither smoking nor alcohol consumption were significantly related to MetS in the CARDIA sample (Carnethon et al., 2004; Duffey et al., 2010), one possible explanation is that those who engage in these behaviors are more likely to drop out of CARDIA. As evidence for this, current smokers accounted for a smaller proportion of the sample at later time points, whereas there was no change in the proportion of former smokers over time. This may indicate a higher dropout rate for those who smoked; had they remained in the study, this might have revealed a relationship between smoking and increased risk of MetS.

For alcohol consumption, it also appears the relationship with metabolic syndrome is more strongly related to drinking high caloric beverages rather than specifically to alcohol use (Duffey et al., 2010). However, the relationship between depression and metabolic syndrome is evident regardless of whether individuals smoke or consume alcohol.

Although not examined in the present study, inflammation has been suggested as a particularly important common biological and bidirectional link between depression and MetS because of its association with both conditions (Capuron et al., 2008; Frank et al., 2021; Sumner et al., 2020). Specifically, studies indicate that an increased inflammatory response caused by a variety of conditions may affect mood and predict depression (Capuron et al., 2008; Frank et al., 2021; Sumner et al., 2020), and that inflammation also increases the likelihood MetS because of the effects of inflammation on of C-reactive protein and interleukin (Capuron et al., 2008; Frank et al., 2021; Sumner et al., 2020). If inflammation can serve as both a causal mechanism and a consequence of both MetS and depression, and this has the potential to create a “vicious cycle” in vulnerable individuals. Specifically, MetS could lead to depression, which in turn, would then worsen the individual’s MetS, and so on.

### Depression and MetS Relations with Sex and Race

In the present study, the relation between depression and MetS was moderated by sex and race (Blacks vs. Whites). Sex independently moderated the relation, but race by itself did not moderate the relation. Furthermore, the relations between depression with MetS were significant for White females, females, and White individuals, but not for other race/sex subgroups. Other studies also support the present results (Beydoun et al., 2020; Cooper et al., 2013; Womack et al., 2016; Yu et al., 2020). Further, the present findings replicate the prior 15-year findings (Womack et al., 2016) from CARDIA over a 30-year period later in the lifespan. One exception is that in the prior study, White males did not demonstrate a significant relationship between depression and MetS. These results may be due to a shorter follow-up, and suggest that some differences emerge in older ages.

It has been suggested that MetS has a “unique phenotype” in Black vs. White adults (Womack et al., 2016). Specifically, this may be partially explained by the fact that dyslipidemia has a significant relation with depression. Black-identified individuals may be less likely to have dyslipidemia as a contributor for MetS, whereas dyslipidemia is often a stronger contributor to MetS in White-identified individuals (Womack et al., 2016). Because dyslipidemia also has a significant relation with depression, it may help to explain the absence of a significant relation between depression and MetS for Black-identified individuals (Womack et al., 2016). As a result, this may show that dyslipidemia is serving as a possible differentiating factor in the variation of the association between MetS and depression along racial lines.

Biological mechanisms alone may not fully explain the differences among race/sex groups in relationships between MetS and depression. There may also be issues with assessment, measurement, and diagnosis of symptoms to identify Black-identified individuals with depression/depression symptomology as there are known variations in presentations along racial lines (Womack et al., 2016). This may most likely manifest in the form of under-reported depression as Black-identified individuals who experience more socioeconomic stress are less likely to report psychological symptoms (Bailey et al., 2019). Furthermore, minority populations are less likely to suffer from acute depression and more likely to suffer from chronic depression than White-identified individuals (Bailey et al., 2019). The higher likelihood that minority populations suffer from chronic depression is a problem that is compounded by the fact that there is evidence to suggest Black-identified individuals are more likely to experience health disparities and access to healthcare issues. (Colen et al., 2018; Noonan et al., 2016).

Also consistent with the present study are prior findings that systemic inflammation might be more pronounced among the subgroup of White-identified individuals (Beydoun et al., 2020). Another study indicates a sex difference in that major depressive disorder is associated with MetS among women but not among men (Yu et al., 2020). This may be due to differences that may exist in the metabolic processes between men and women. However, this may also be due to the ways in depression is reported, recorded, and treatment is sought out which may be different across sex lines similar to differences seen in race (Yu et al., 2020).

It should be noted that there are some contradictory findings that indicate a link between depression and MetS among those identified as Black, particularly Black women (Cooper et al., 2013). In contrast to our findings, the latter study found that hypertension, obesity, and HDL-C contributed most to the link between depression and MetS (Cooper et al., 2013) whereas we did not find an association between depression and either hypertension or obesity. Since causal factors (i.e., hypertension and obesity) were not related to depression MetS itself would not be significant for this sample either.

### Study Limitations and Strengths

Although this study is prospective, a limitation is that other than examining health behaviors, it did not evaluate possible causal mechanisms or explanatory links for the relation between depression and MetS. There are also limits to the generalizability of the findings from this study. The study cohort included individuals who identified as White or Black, but the study did not include other racial or ethnic groups. Additionally, this study’s sample population was aged 18-30 at intake and 48-60 at the latest available timepoint, so the results may not be generalizable to other age groups. There may also be limitations to these conclusions, as the CESD measure for depression may not be sensitive enough to identify clinical depression, and may be subject to racial differences in depression symptoms (Bailey et al., 2019; Gurka et al., 2014; Womack et al., 2016). Results may also be specific to use of the NCEP-ATP III criteria to define MetS (Huang, 2009), which requires that individuals subjects must meet three of five components for MetS.

There are several notable strengths of the study. The CARDIA study cohort allowed us to examine relationships between depression and MetS in a bi-racial cohort with equal numbers of males and females allowing for a robust test of race and sex as moderators. Furthermore, this study looked at a cohort over the course of 30 years comparing longitudinal and cross-sectional data across those 30 years. Additionally, because of the CARDIA study design with multiple assessments of both variables, we were able to evaluate predictive bi-directional relationships between depression and MetS in the same cohort using lagged time series analyses.

The present findings have several implications for future research on depression, MetS, and relations between the two. First, having established a bidirectional relation between depression and metabolic health in the same cohort, there is a need to determine mechanisms that might mediate these effects in both directions. These potential causal mechanisms may be evaluated in a future study by looking evaluating a potential mediator such as cortisol or inflammatory cytokines. Another issue needing investigation is whether these effects are evident in individuals over the age of 60 years. To better understand the moderating effects of sex and race, further research should determine whether differences are evident with other measures of depression (e.g., clinical interview or clinician ratings) that are not solely based on self-report and that may be less subject to biases due to sex and race. In addition, examining the underlying mechanisms of how MetS and depression are evaluated and assessed across race and sex lines may allow for better diagnosis of conditions in different populations.

## Data Availability

The National Heart, Lung, and Blood Institute distributes these data as a publicly available dataset from the NHLBI CARDIA Study. Procedures for obtaining these data are found at https://biolincc.nhlbi.nih.gov/home/

